# Vaccination of solid organ transplant recipients previously infected with SARS-CoV2 induces potent responses that extend to variants, including Omicron

**DOI:** 10.1101/2022.02.10.22270607

**Authors:** Alok Choudhary, Mark Lerman, David Calianese, Salman Khan, Judson Hunt, Afzal Nikaein, Avi Z. Rosenberg, Jonathan I. Silverberg, Israel Zyskind, William Honnen, Dabbu K. Jaijyan, Erica Kalu, Abraham Pinter

## Abstract

**Background:** Multiple factors affecting COVID19 vaccine induced antibody responses in SARS-CoV2 uninfected immunosuppressed solid organ transplant recipients have been reported; however, there is still a lack of information on non-ACE2 competing cross-CoV2 neutralizing functional antibodies induced in these cohorts, and similarly the vaccine efficacy in prior CoV2-infected immunosuppressed individuals is not well understood.

**Methods:** COVID19 vaccine efficacy was compared in a panel of kidney and heart transplant recipients who were either CoV2 uninfected (n=63) or CoV2 infected (n=13) prior to receiving two or three doses of mRNA vaccines using pseudoviral neutralization assays against eight CoV2 strains (the CoV2_D614G ancestral strain, alpha, beta, gamma, delta, kappa, lambda, and omicron-BA1 variants), while plasma antibody titers were determined by ELISA using recombinant CoV2-RBD-wt proteins.

**Results:** Minimally protective neutralizing plasma antibody titers (IC_50_ ≥ 1:50) against the variants were recorded 7-14% and 25-35% after the second and third doses respectively, with Omicron being the most resistant. In contrast, all previously infected vaccinees possessed minimal protective plasma titers against D614G after either two or three vaccine doses, with 11/13 exhibiting strong protection (IC50≥ 1:500) and 10/13 exceeding the minimal protective titer against Omicron. Absorption of the selected plasma with immobilized parental RBD removed ≥ 90% of its neutralizing activity, indicating that the dominant neutralization targets were in the RBD.

**Conclusions:** This study showed that CoV2 infection followed by vaccination, but not vaccination alone, induces the presence of potent highly cross-reactive CoV2 neutralizing plasma antibodies that extend to Omicron variants, even in immunosuppressed SOTRs.

## INTRODUCTION

As of early July 2022, the cumulative number of confirmed infections caused globally by SARS-CoV-2 (CoV2) is approaching 546 million, with close to 6.3 million deaths, whereas the US alone has reported more than 87 million cases, with ≥ 1 million deaths^**1**^. While the rapid development of various COVID-19 vaccines has helped control the spread of this virus and greatly reduce severe complications^**2-8**^, the emergence of more infectious and resistant CoV2 variants has resulted in spikes in the number of infections and deaths, which are centered on unvaccinated populations and people with impaired immune responses, including solid organ transplant recipients (SOTRs)^**1,9-15**^.

Several reports have indicated that most SOTRs possess limited immunity after the standard two-dose regimen of RNA-based vaccines^**16-19**^,whereas a third dose of the vaccine results in increased titers of antibodies targeting the receptor-binding domain (RBD) and other subdomains of the CoV2 spike protein^**20-27**^.A recent study evaluating the effect of a fourth dose of COVID-19 vaccines in 18 SOTRs showed positive antibody responses in subjects who were either negative or very low positive prior to receiving the latest boost^**28**^.

Mounting evidence indicates that sites in the RBD of the CoV2 spike protein contain major targets of protection in COVID19 convalescent and healthy vaccinated individuals^**29-31**^. The dominant neutralizing response is directed against sites in the receptor-binding motif (RBM) of the RBD, which mediates recognition of the Ace2 receptor^**32**^. Unfortunately, these targets are highly mutated in the recently emerged Omicron variant and its subvariants BA1, BA1.1, BA2, BA4, or BA5, and antibodies directed against this site bind poorly and fail to neutralize these viruses^**33,34**^. With the increasing spread and dominance of Omicron-related variants, this raises concerns about the efficacy of current vaccines^**35-39**^, particularly in this population.

To explore the COVID-19 vaccine-induced immunity in SOTRs, we used RBD-specific binding^**40**^ and absorption assays to quantify and determine the specificity of protective immune responses in a cohort of 76 heart and kidney transplant patients after their second (n=51) or third (n=25) COVID-19 vaccine dose. Plasma neutralizing titers were determined using pseudotyped viruses expressing the D614G parental strain and alpha, beta, gamma, delta, kappa, lambda, or omicron variant spikes^**41**^. For several strongly neutralizing plasma samples, the specificity of this protective activity against RBD was analyzed by absorption with immobilized recombinant RBD proteins.

The results of this study indicate that a relatively strong neutralizing response to the parental strain and many CoV2 variants was induced in SOTRs who were CoV2-infected and vaccinated, whereas SOTRs who were not previously infected with the virus exhibited weak or undetectable neutralizing responses. This cross-reactive neutralizing response was also observed to a lesser extent for omicron variant. Several factors associated with reduced protective response have been identified. A better understanding of the mechanistic basis of this cross-neutralizing response may allow enhanced vaccine strategies and responses in these populations in the future.

## MATERIALS AND METHODS

### Study design and patient population

This study was aimed at recruiting kidney and heart transplant patients who had been vaccinated with two or three doses of a COVID-19 vaccine manufactured by Pfizer or Moderna or a single dose of the J&J vaccine to investigate the demographic and clinical factors behind the poor immune response to COVID-19 vaccines in SOTRs. Kidney transplant recipients, heart transplant recipients, or kidney+heart transplant recipients were recruited after their second vaccine dose (n=51) during routine visits to the transplant clinic at Medical City Hospital, Dallas between June 22 and July 22, 2021, and after their third vaccine dose (n=25) between Nov 8 and Dec 21, 2021. All the participants provided written informed consent. The majority of the patients did not report previous CoV2 infections, but ∼15% of the post-second dose and ∼20% of the post-third dose cohort had PCR-confirmed CoV2 infections prior to vaccination. Considering the requirement of a five-month interval for booster shots^**42**^, the blood samples were collected up to five months away from their 2^nd^ and 3^rd^ vaccine shots. Blood samples were collected in sodium-heparin tubes and processed for plasma separation by centrifugation. Plasmas were aliquoted in multiple 1 ml vials and stored at -80°C for subsequent analyses of antibody binding and CoV2 neutralization activity.

### Expression and purification of the CoV2 receptor-binding domain (RBD) protein

RBD-wt antigens were expressed as fusion proteins with the gp70 carrier domain, as previously described^**43**^. In brief, a gene fragment of the CoV2-Spike gene encoding the RBD was synthesized commercially (Integrated DNA Technologies, Coralville, IA) and cloned at the 3’ end of a gene expressing the N-terminal fragment of the Friend ectotropic MuLV (Fr-MuLV) surface protein (SU) gp70 gene in the expression vector pcDNA3.4 (Addgene, Watertown, MA). The resulting plasmid was transfected into 293F cells using the Expi293 Expression System (Thermo Fisher Scientific, Waltham, MA), according to the manufacturer’s protocol.

### RBD plasma antibody detection by enzyme-linked immunosorbent assay (ELISA)

RBD plasma antibodies were detected using an RBD-wt protein-binding ELISA as described earlier^**43**^. In brief, RBD-wt protein was coated overnight at 4°C at a concentration of 100 ng/well in 50 µl of bicarbonate buffer (pH=9.8) using U-shaped medium binding 96-well ELISA plates (Greiner Bio-One; Cat#:650001). 2% nonfat dry milk was used to make the plasma dilution and to block the plate. Plasma from vaccinated SOTRs was prepared at different dilutions (1:2, 1:10, 1:50, 1:250, 1:1250, 1:6250) and incubated at 50 µl/well in duplicates for one hour at 37°C in a blocked plate after washing with wash buffer (1XPBS+0.1% Tween 20) and Alkaline phosphatase conjugated goat anti-human IgG, IgM and IgA detector antibodies (*Jackson ImmunoResearch* Laboratories, West Grove, Pa) were used to detect the plasma RBD-IgG, IgM and IgA. The quantitative value of plasma RBD antibodies was calculated using the area under the curve (AUC) after plotting values from all different dilutions of plasma using Graph Pad Prism 8.0.

### CoV2 pseudovirus (psV) preparation and ACE2-HeLa cell-based neutralization Assay

Codon-optimized D614G, alpha, beta, gamma, delta, kappa, lambda, and omicron spike gene sequences with 18 aa C-terminal truncations were cloned into the pCAGGS vector^**29**^. Instead of the D614D Wuhan ancestral strain^**44**^,which is also a COVID-19 vaccine strain, D614G was used because this mutant had higher transmissibility, which led to the pandemic, and had no difference in immunogenicity^**45,46**^. D614G and seven different CoV2 pseudovirus variants were generated by co-transfecting spike and pnl4.3. Luc.r^-^e^-^ plasmids into HEK 293T cells to produce the CoV2 psV as described earlier^**41**^ and HuACE2-HeLa cells were infected with the CoV2 psV in DMEM media supplemented with polybrene (10 µg/ml), and infectivity was determined by reading luciferase activity in the cell after 72 h post-infection upon adding luciferase substrate (Britelite, PE) to the cell-lysate. To determine the neutralization potency of plasma psV dilutions, ∼100,000 RLU was used to infect Hu-ACE2-HeLA cells in the presence of titrated plasma.

### Preparation of CNBr-RBD Sepharose bead column and absorption of plasma RBD antibodies

Cyanogen bromide (CNBr)-activated Sepharose 4 B(Pharmacia#17-0430-01) beads after hydration with 1mM HCl for 30 min at room temperature were used for conjugation with RBD-wt CoV2 protein in 1X PBS in a tube by incubating overnight at 4°C on a vertical rotator. After washing with excess 1X PBS RBD-conjugated beads were blocked with 1M ethanolamine (pH 8.0) for two hours at room temperature on a vertical rotator. Blocked beads were washed and were stored in the refrigerator for plasma RBD antibody absorption experiments.

1000 ul of 10-fold diluted plasma from selected patients were incubated O/N at 4°C on vertical rotator. Tubes with different plasma samples were spun at 300 x g to collect the supernatant as the absorbed plasma RBD antibody fractions. Absorbed and unabsorbed plasma at the same dilutions were tested for RBD-wt and NTD binding by ELISA to determine the percentage absorption and change in unabsorbed plasma antibody concentrations. Absorbed RBD-Plasma Ab fractions were stored in -80°C for neutralization experiments together with the unabsorbed plasma.

### Statistical Analysis

GraphPad Prism (version 8.0) was used to calculate the mean, median, and interquartile range (IQR), and to determine suitable parametric or nonparametric tests for statistical analyses of the data. Unpaired Student’s t-tests or Mann-Whitney tests were performed for column statistical analysis to compare two different groups whereas paired t-tests were performed to compare the neutralization potency of plasma against multiple CoV2 variants. Pearson’s coefficient and Spearman’s r values were calculated to identify correlations. The neutralization potency of plasma, defined as the plasma dilution, required to reduce viral infection by 50% (IC50), was calculated using One-Site Fit LogIC_50_ regression. P-values were calculated at a confidence interval of 95% and are indicated as <0.05, or *; <0.01, or **; and <0.001, or ***. An IC_50_ ≥ 1:50 was considered above the minimal protective dose, as determined by antibody transfer protection experiments in macaques^**47**^.

## RESULTS

### COVID-19 vaccinated SOTRs with prior CoV2 infections had stronger CoV2-RBD antibody responses than uninfected vaccinated subjects

A description of the cohort tested in this study is provided in **Table 1**, including the distribution of age, sex, race, organ transplant, immunosuppressive regimens administered, vaccine types, and days post-vaccination. Of the 76 SOTRs, 47 received two doses of the mRNA CoV2 vaccine, 25 received three doses of the same vaccine, and four SOTRs had received a single dose of the J&J vaccine.

**Table. 1.** Demographic and clinical characteristics of the 76 SOT recipients

|  | Post-second | Post-third |
| --- | --- | --- |
|  | Dose | Dose |
| **Total number, n** | 51 | 25 |
| **Age (Mean ± SD)** | 59 ±15 | 64 ±10 |
| Age group 18-59, n (%) | 21 (41%) | 6 (24%) |
| ≥60, n (%) | 30 (59%) | 19 (76%) |
| **Sex** |  |  |
| Female gender, n (%) | 27 (53%) | 10 (40%) |
| Male gender, n (%) | 24 (47%) | 15 (60%) |
| **Race** |  |  |
| Black (African American), n (%) | 16 (31%) | 7 (28%) |
| Hispanic, n (%) | 8 (16%) | 4 (16%) |
| White, n (%) | 26 (51%) | 13 (52%) |
| Asian, n (%) |  | 1 (4%) |
| **Type of Organ Transplantation** |  |  |
| Heart, n (%) | 19 (37%) | 13 (52%) |
| Kidney, n (%) | 29 (57%) | 11 (44%) |
| Heart and Kidney, n (%) | 3 (6%) | 1 (4%) |
| **Time since Transplantation (years)** |  |  |
| 0 to 1.5 | 24 | 12 |
| >1.5 | 27 | 13 |
| **Type of maintenance immunosuppression** |  |  |
| Sirolimus /Tacrolimus+Myfortic+Prednisone | 31 | 12 |
| Sirolimus/Tacrolimus+Myfortic | 13 | 10 |
| Sirolimus/Tacrolimus | 0 | 1 |
| Tacrolimus/Prednisone | 3 | 0 |
| Myfortic/Prednisone/Belatacept | 2 | 1 |
| Prednisone/Cyclosporin | 1 | 0 |
| Myfortic/Cyclosporin | 0 | 1 |
| Sirolimus/Tacrolimus/Myfortic/Prednisone/Cs | 1 | 0 |
| **CoV2 Infection Status** |  |  |
| CoV2 Uninfected + Vaccinated, n (%) | 43 (84%) | 20 (80%) |
| CoV2 Infected + Vaccinated, n (%) | 8 (16%) | 5 (20%) |
| **Type of COVID19 Vaccine** |  |  |
| Pfizer, n (%) | 34 (67%) | 17 (67%) |
| Moderna, n (%) | 13 (25%) | 8 (25%) |
| J&J, n (%) | 4 (8%) | 0 |
| **Postvaccination time (Months)** |  |  |
| < 2, n (%) | 15 (30%) | 7 (30%) |
| 2- 3, n (%) | 16 (31%) | 12 (31%) |
| 3-5, n (%) | 20 (39%) | 6 (39%) |

We used a novel RBD-gp70 fusion protein system to express native RBD as an antibody target, which recognizes virus-specific antibodies more efficiently than the standard RBD antigens^**43**^. All 76 SOT recipient’s plasmas were screened for levels of anti-CoV2 -RBD IgG, IgM, and IgA antibodies and 3X background value with pre-pandemic healthy plasma RBD binding (AUC of 10) was considered as cut-off for positive RBD antibody titer.

SOTRs have a lower immunological response to influenza vaccination during the first six months after transplantation^**48**^. Consistent with earlier findings, we observed relatively weak antibody response to COVID19 mRNA vaccines in SOTRs who were vaccinated less than 1.5 Y from their transplantation, whereas those SOTRs vaccinated later than 1.5 Y after transplantation had significantly better anti-RBD-IgG responses (**Fig.1A**).

**Fig. 1.**
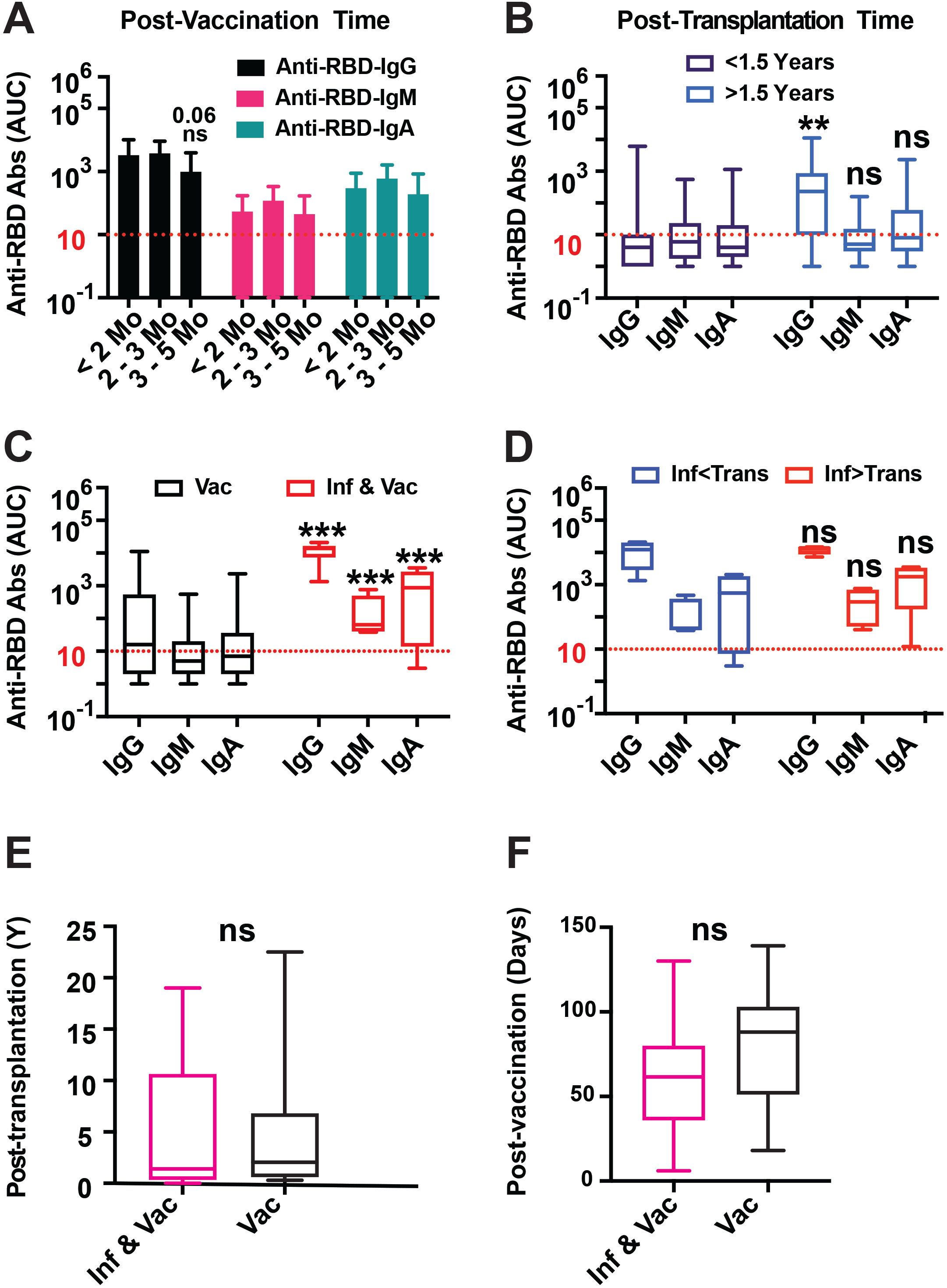
Post-vaccination anti-CoV2-RBD plasma IgG, IgM, and IgA response in SOTRs. **(A)** Comparison of anti-RBD plasma IgG, IgM, and IgA levels among three different groups of SOTRs with <2 months, 2-3 months and ≥ 3 postvaccination time. **(B)** Comparison of anti-RBD plasma IgG, IgM, and IgA levels between group of SOTRs vaccinated within or after 1.5 Y since kidney or heart transplant. **(C)** Comparison of vaccine induced RBD-antibodies isotypes between CoV2 infected-vaccinated and CoV2 uninfected-vaccinated SOTRs. **(D)** Anti-RBD plasma IgG, IgM, and IgA levels between group of infected-vaccinated SOTRs who reported CoV2 infection prior to transplantation vs after transplantation. **(E)** Comparison of post-transplantation time in Y between CoV2 infected-vaccinated and uninfected-vaccinated SOTRs **(F)** Comparison of post-vaccination time in days between CoV2 infected-vaccinated and uninfected-vaccinated SOTRs. Horizontal red dotted lines represent CoV2 RBD antibody positive cut-off value.

Because of the waning immunity of the COVID19 vaccine over time, a booster dose is recommended to achieve effective protection^**49-50**^. As expected, around 40% of the total participants who were recruited between 3-5 months after their second and third dose showed lower anti-RBD IgG titers compared to those who were recruited within 3 months after vaccination (**Fig.1B**).

Most vaccinees receiving two or three mRNA vaccines doses (63 of 76) reported no previous CoV2 infections, whereas 13 of 76 reported prior CoV2 infections as confirmed by PCR (**Table 1**). Approximately half of the vaccinees (5/13) were infected before transplantation and the remainder were infected after transplantation. There were significantly higher levels of all three anti-RBD immunoglobulin isotypes in the infected vaccinees than in the uninfected vaccinees, whereas RBD antibody titers were comparable between vaccinated SOTRs who reported infections before transplantation and those who were infected after transplantation (**Fig.1C**). In contrast to the CoV2 uninfected SOTRs, no significant differences were observed in the anti-RBD plasma antibody titers of the CoV2-infected SOTRs vaccinated before or later than 1.5 Y of transplantation (**Fig.1D**).

### COVID-19 vaccinated SOTRs with prior CoV2 infection possessed considerably higher cross-CoV2 neutralizing plasma antibody titers

All 76 SOTRs plasma samples were assayed for their neutralization potency in an HuACE2-HeLA cell-based assay against pseudoviruses containing eight different spike proteins. These included the parental D614G spike, two different variants of interest (VOI), and five variants of concern (VOC), including a representative of the currently circulating highly contagious and resistant omicron subvariant BA1^**51**^. Among the uninfected SOTRs vaccinees only 4/63 showed strong neutralization potency (IC50s ≥ 1:500) against D614G, and 16/63 possessed neutralizing titers above the minimal protective titer (IC50≥ 1:50) against D614G. In contrast, 11/13 of the plasma samples from previously infected vaccinees possessed IC50s≥ 1:500, and all 13 of these vaccinees had titers above the minimal protective level for the D614G pseudotype (**Fig. 2A**).

**Fig. 2.**
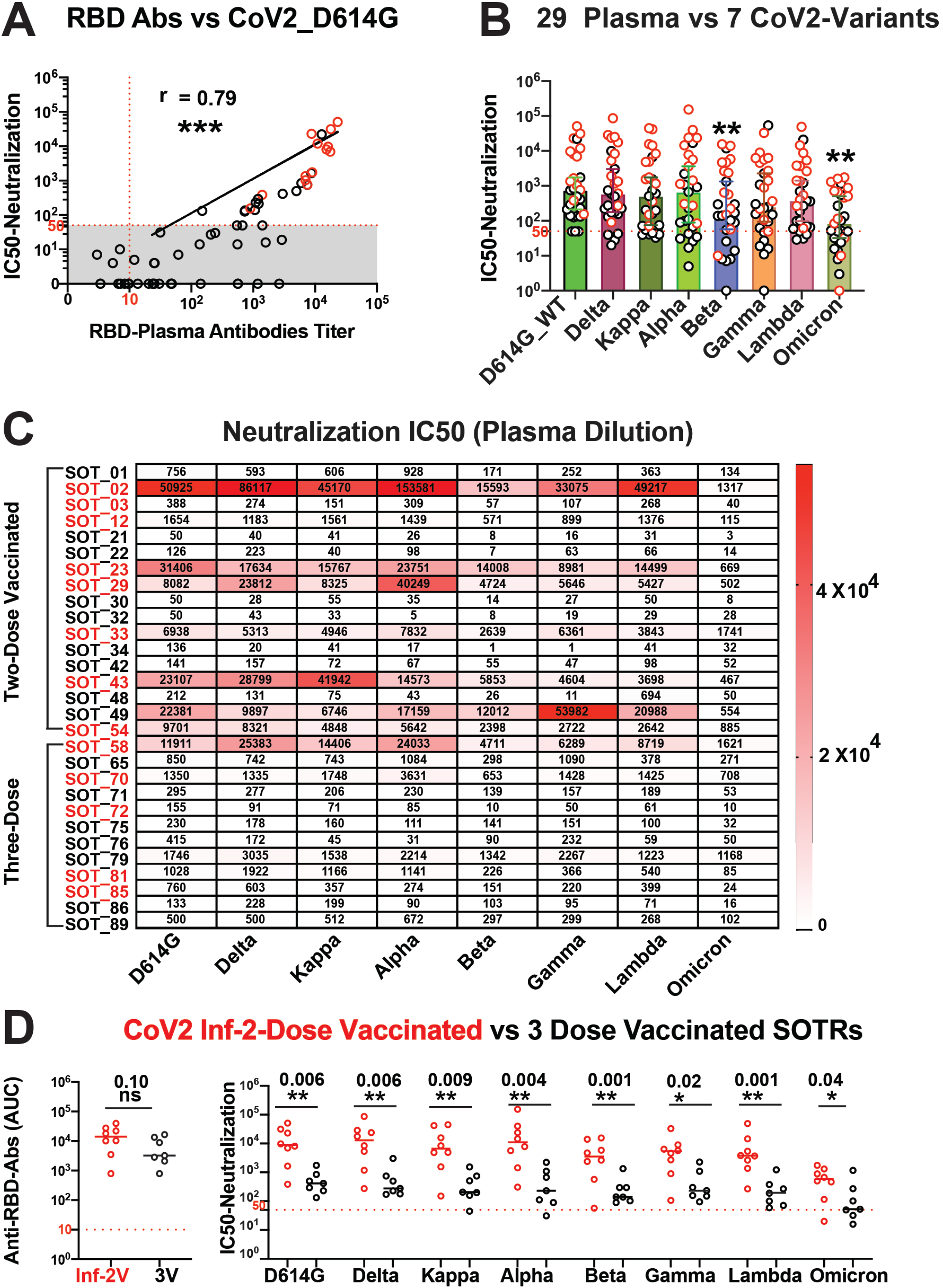
Plasma RBD antibodies titers correlate with CoV2 variant neutralization potency. **(A)** Correlation between neutralization potency (IC50) of SOTRs plasma ((infected/vaccinated (**O**) and uninfected/vaccinated (**O**)) against D614G and their CoV2-wt RBD antibodies titer **(B)** Neutralization potency of 29 selected plasma with IC50≥1:50 against D614G (16 uninfected-vaccinated and 13 infected-vaccinated) against 7 different variants including BA1 Omicron subvariant. **(C)** Neutralization IC50 values of 29 selected SOTRs’ plasma against 8 different CoV2 including baseline D614G(**D**) Comparison of anti-RBD-wt plasma antibody binding titer and neutralization potency against CoV2 variants between CoV2 infected two doses vaccinated (Inf-2V) and CoV2 uninfected three doses vaccinated (3V) SOTRs. Horizontal red dotted line represents minimal required protective IC50 of plasma for virus neutralization (1:50 dilution) and cut-off for the RBD positive serology (AUC of 10).

All 29 plasma samples (13 infected and 16 uninfected vaccinees) with minimal protective titers against D614G were further tested against each of the eight CoV2-variants to compare their cross-CoV2 neutralization potencies. Comparable neutralization titers were observed against D614G, alpha, gamma, lambda, and delta variants in all plasma samples. A significant decrease in neutralization potency was observed for all twenty-nine plasmas for the beta variant, and an ever-greater decrease (∼10 fold lower) was observed for most of the plasma for omicron compared to the other viruses (**Fig. 2B**). The greater resistance of the omicron virus is consistent with that reported for healthy vaccinees^**35,52-54**^. Despite this decreased potency, most (7/13) of the infected vaccinee plasma retained strong neutralization titers for the omicron pseudotype (IC50 ≥ 1:500), compared to only 3% (2/63) of the uninfected vaccinee plasma with this level of activity (**Fig. 2C**).

In the two-dose-vaccinated CoV2 uninfected cohort, only 21% (9/43) of SOTRs reached the minimal protective titer (IC50≥ 50) against D614G, which decreased to 9% (4/43) against the highly resistant omicron variant. In CoV2 uninfected cohorts, three-dose-vaccinated SOTRs showed improved immunity compared to the two-dose-vaccinated SOTRs, with 35% of SOTRs reaching the minimal protective titer against D614G and 15% against Omicron (**Fig.2C**).

Considering that CoV2-infection is equivalent to an additional dose of vaccine, we compared the CoV2-infected-2-dose vaccinated SOTRs with CoV2-uninfected-3-dose vaccinated subjects with IC50 ≥ 1:50. Interestingly, we noticed no significant difference in the plasma RBD antibody titers between these two groups, but a significantly reduced neutralization potency was observed in the CoV2-uninfected 3-dose-vaccinated group against all the tested variants as compared to CoV2-infected-2-dose vaccinated individuals (**Fig.2D**).

### Evidence that vaccination boosted pre-existing antibody specificity against conserved neutralization targets in CoV2 infected SOTRs

Due to the unavailability of pre-vaccination plasma for infected SOTRs vaccinees, we were unable to determine the relative contributions of infection versus vaccination to the cross-reactive and Omicron-specific neutralization responses of these subjects. We indirectly approached this question by comparing the plasma neutralization potencies of CoV2 convalescent plasma from healthy subjects collected early in the pandemic. Like most CoV2 infected SOTRs, these individuals were reported to have been infected before July 2020 before the availability of CoV2 vaccines. The convalescent plasma contained comparable neutralization potency against the parental D614G virus, but considerably lower neutralization potencies for the Omicron variant than the infected SOTR vaccinees. This suggests a critical role of vaccination in the induction of protective cross-reactive antibodies that target conserved neutralizing epitopes present in the RBD outside the immunologically dominant RBM region in immunosuppressed SOTRs infected with CoV2 prior to vaccination (**Fig. 3A**).

**Fig. 3.**
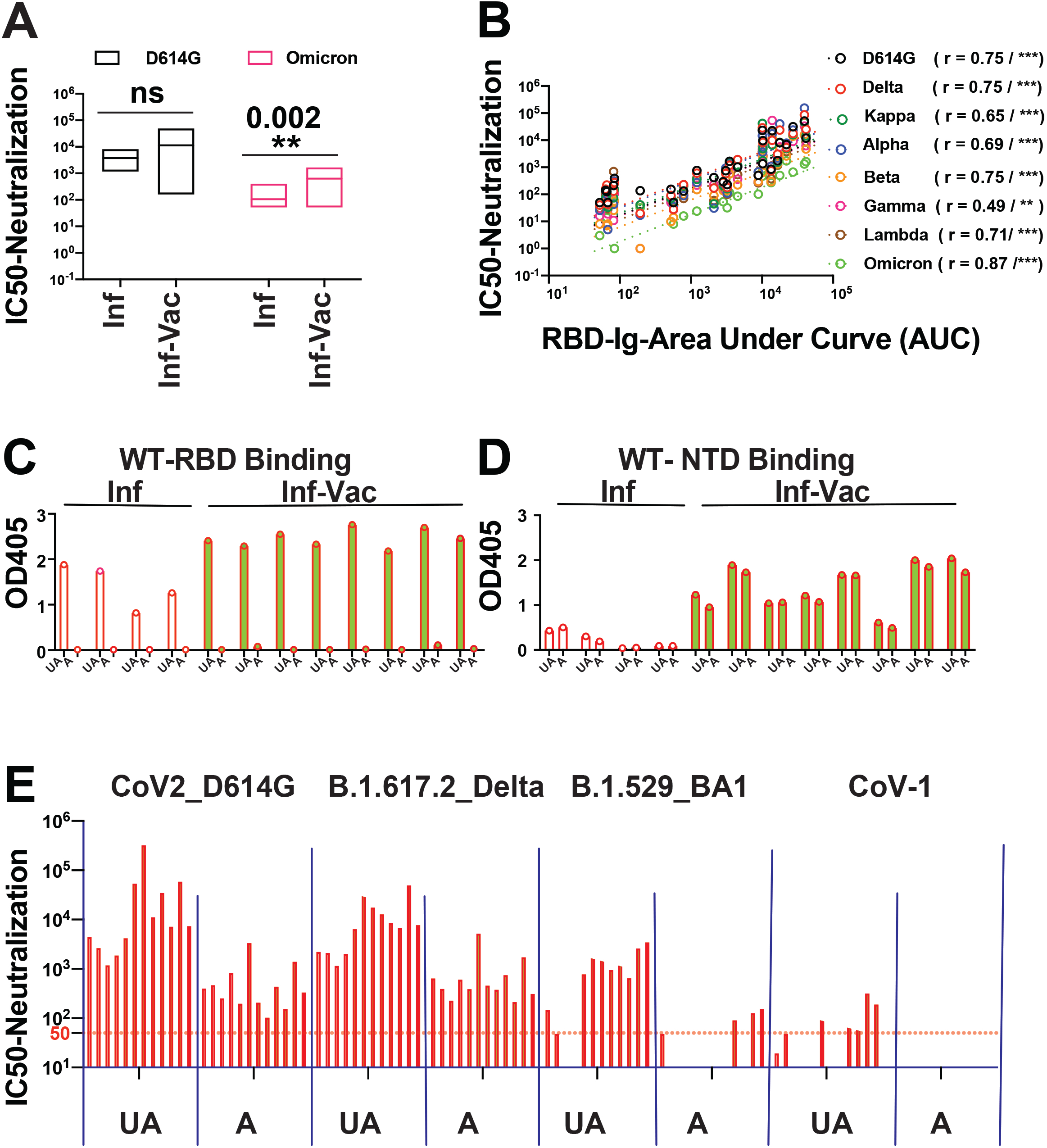
Induction of antibodies against the conserved RBD neutralizing epitopes are responsible for the cross-CoV2 neutralization potency of infected-vaccinated SOTRs’ plasma. **(A)** Comparison of D614G and BA1-Omicron neutralization potency between CoV2-infected COVID19-unvaccinated healthy subjects and CoV2-infected COVID19-vaccinated SOTRs **(B)** Correlation between anti-RBD-wt plasma antibody titer and CoV2 variants neutralization potency for 29 selected plasma with IC50≥1:50 against D614G **(D)** Binding of 12 selected RBD-wt absorbed and corresponding unabsorbed plasma with RBD-wt to determine the % antibody RBD-wt antibody absorption **(E))** Binding of 12 selected RBD-wt absorbed and corresponding unabsorbed plasma with NTD-wt to determine the % non-RBD-wt antibody loss. **(F)** Neutralization potency of 12 selected RBD-wt absorbed and corresponding unabsorbed plasma against D614G, Delta, BA1-Omicron and CoV1. Horizontal red dotted line represents minimal required protective IC50 of plasma for virus neutralization (1:50 dilution).

As expected from previous studies, strong correlations were observed between plasma RBD antibody titers and neutralization titers against D614G-wt and all the CoV2 variants (**Fig. 3B**) The importance of epitopes in the RBD as neutralization targets in these plasma samples was tested by examining the effect of RBD-specific antibody depletion from the selected plasma on neutralization potency and specificity.

Selected plasma Ab from CoV2-Infected-unvaccinated and CoV2-infected-vaccinated individuals were absorbed by passage through wt-RBD-conjugated CnBr beads. The column flow-through was tested for CoV2-wt-RBD and N-terminal domain (NTD) binding by ELISA, and for neutralization activities against D614G, delta, omicron-BA1, and CoV-1 pseudoviruses. Using ELISA, we showed that passage over these columns absorbed almost 100% of wt-RBD binding activity (**Fig. 3C**), whereas the NTD-specific antibodies remained unaffected (**Fig. 3D**). A low level of residual neutralizing activity (<10%) was observed in the column flow throughs for the D614G and delta variants, but not for the Omicron and CoV1 pseudotypes (**Fig. 3E**). This experiment confirms the dominant role of conserved sites in the RBD as neutralization targets, and further suggests that conserved neutralizing epitopes outside of the highly mutated receptor-binding motif (RBM) were responsible for the neutralization activity against the Omicron variant.

### Prior CoV2 infection led to an optimum immune response in SOTRs of African American descent and in those who were vaccinated close to their date of transplantation

Inherent differences in the immune system have been shown to be responsible for the disproportionate mortality rate in the black population due to COVID19^55^. Race and ethnicity are known to affect the antibody responses to rubella and influenza vaccines, which significantly induces higher titers in those of African American ethnicity as compared to those of White or Hispanic^**56,57**^.

To examine whether the COVID-19 vaccine-induced antibody response in CoV2 uninfected vaccinated cohorts was affected by race, we compared RBD antibody titers in three different ethnic groups: Black, Hispanic, and White. Contrary to the findings for rubella and influenza vaccines, we noticed no significant difference in plasma RBD antibody levels between Hispanic and white participants. However, SOTRs of African American descent had significantly lower RBD antibody titers than those identified as white (p=0.003) or Hispanic (p=0.013) (**Fig.4A**).

**Fig. 4.**
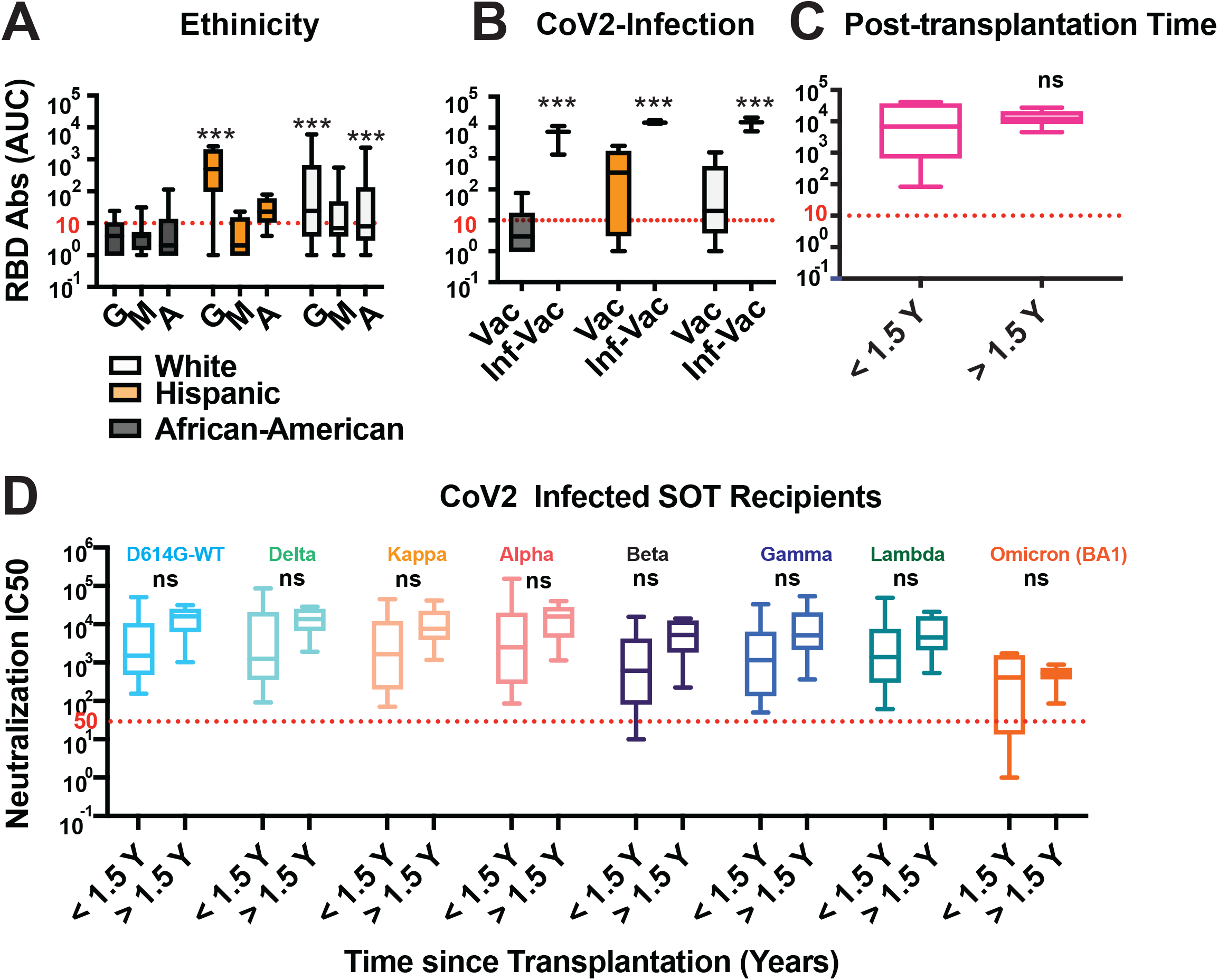
SOTRs of African American ethnicity and those who were vaccinated close to their transplant time and history of CoV2 infection showed optimal response to vaccine as compared to their corresponding group with no CoV2 infection. **(A)** Comparison of vaccine induced anti-RBD-IgG, IgM, and IgA in CoV2 uninfected SOTR of different ethnic groups. **(B)** Anti-RBD-wt antibody plasma titer between CoV2 infected-vaccinated vs uninfected-vaccinated SOTRs of same ethnic group **(C)** Comparison of anti-RBD plasma antibodies titers in CoV2 infected SOTRs who were vaccinated within 1.5 Y of transplantation and those who were vaccinated after 1.5 Y of transplantation **(D)** Effect of serum creatinine level (mg/dl) on the vaccine induced antibody response. **(E)** Comparison of CoV2 variants neutralization potency between CoV2 infected SOTRs who were vaccinated within 1.5 Y of transplantation and those who were vaccinated after 1.5 Y of transplantation. Horizontal red dotted line represents minimal required protective IC50 of plasma for virus neutralization (1:50 dilution) and cut-off for the RBD positive serology (AUC of 10).

(To our surprise) none of the CoV2 infected-vaccinated SOTRs of African-American ethnicity had a weak immune response to the COVID-19 vaccine (**Fig.4B**). A similar observation was made for SOTRs vaccinated close to their transplantation date when they were on aggressive immunosuppressive drug regimens. In contrary to CoV2 uninfected SOTRs, CoV2-infected SOTRs vaccinated within 1.5 Y of transplantation showed strong anti-RBD antibody response (**Fig.4C**) and the neutralization potencies were also comparable between the groups vaccinated before or after 1.5 Y since their transplants (**Fig. 4D**). These studies showed that CoV2 exposure before vaccination had a dramatic effect on the induction of antibodies against cross-CoV2 conserved neutralizing epitopes, even in the presence of an immunosuppressive environment.

## DISCUSSION

Immunosuppressed SOTRs have been reported to have poorer responses to Covid-19 vaccines than healthy populations, with one early study (before the advent of omicron variants) showing that SOTRs had an 82-fold higher risk of breakthrough infection and 485-fold higher risks of associated hospitalization and death compared to the general population^**58,59**^.

Several previous studies on SOTR responses have used simple binding assays or surrogate virus neutralization tests (sVNT) based on ACE2 binding competition to measure neutralizing antibody responses^**22,28,60-65**^.

However, only a fraction of CoV2 spike or RBD-binding antibodies possess neutralization activity^**66-68**^; therefore, binding assays are not good correlates for protection. Many of the Omicron-neutralizing mAbs target conserved regions of the RBD core outside of the RBM, or sites in the N-terminal domain (NTD) that are not involved in ACE2 competition^**69-72**^, and prevent infection by an alternate mechanism that interferes with Ace2 binding^**73**^. To avoid these limitations, the present study used both binding assays and direct measurements of variant-specific neutralization titers to accurately evaluate antibody-mediated protection of SOTRs.

The most striking observation in this study was the significantly higher neutralization titers of SOTRs who reported CoV2-infections prior to vaccination than those who were not previously infected (**Fig.1C**). Previously infected subjects possessed higher RBD antibody binding titers, higher neutralization potencies, and greater neutralization breadths than fully vaccinated uninfected SOTRs, including those who received an additional booster vaccination. Of particular interest, neutralization titers of infected-vaccinated SOT recipients were not only greater than those of triply vaccinated SOT recipients but were also significantly enhanced against the highly mutated Omicron strain.

RBD absorption experiments showed that the dominant specificity of this activity was directed against conserved epitopes shared by Wuhan and Omicron sequences. Although pre-vaccination bleeds were not available in this study, the known short half-life of the neutralizing response following CoV-2 infection and the considerable amount of time elapsed since infection in most SOTRs suggest that the initial response to infection should have been significantly diluted. In addition, control sera from healthy subjects infected early during the pandemic, before vaccines were available, selectively neutralized the D614G strain compared to omicron BA-1 (**Fig. 3A**), confirming other reports that neutralizing antibodies induced by infection with early strains strongly prefer the wt sequence over the omicron variant. These results indicated that it is highly likely that vaccination following infection was responsible for the enhanced responses observed in these subjects.

Interestingly, breakthrough omicron infections in vaccinated healthy individuals have also been reported to induce cross-CoV2 neutralizing antibodies that target conserved neutralizing epitopes in RBD^**74**^. This can be explained by the fact that the omicron spike cannot restimulate memory B cells specific for the dominant and highly immunogenic epitopes present in the RBM, because of the high level of RBM mutations in these variants. This reduced competition allows for the stimulation of cells that recognize the less immunogenic subdominant neutralizing epitopes conserved in the RBD and elsewhere in the spike antigen. It is unclear why vaccines based on the Wuhan strain also induce cross-neutralizing antibodies against highly conserved sub-dominant targets in subjects previously infected with the original strain or early variants but not in uninfected subjects vaccinated with the Wuhan strain. A possible explanation is that infection is more efficient than current vaccination strategies for inducing B cell recognition of subdominant targets in the spike antigen, perhaps due to more accurate presentation of native structures and conformations, or better T cell help due to strong germinal center reactions, possibly stimulated by epitopes encoded by other genes of the virus, as has been previously suggested for stronger antibody responses in healthy individuals with prior Covid19 infections^**75-77**^,

An important factor identified in this study that inhibited an effective neutralizing response was the length of time elapsed since the transplantation. It has been previously reported that a longer post-transplantation time positively correlates with the efficacy of vaccine-induced humoral responses in SOTRs^**62**^. In our study, quantitation of plasma RBD antibody titers allowed the delineation of the post-transplantation time point within which SOTRs were more likely to have weak antibody responses to the vaccine. In contrast to earlier findings on influenza vaccines that 6 months post-transplantation was the optimal time for immunogenicity, we found that 18 months (1.5 Y) post-transplantation was a better predictor of a stronger antibody response to the Covid19 vaccine in SOTRs. We showed that prior infection increased vaccine efficacy, even in SOTRs who were vaccinated close to their transplantation time and were supposedly on a more aggressive drug regimen for maintenance immunosuppression. Consistent with earlier observations^**78**^, we found weaker antibody responses to vaccines in CoV2 uninfected African-American SOTRs, which was not consistently observed in CoV2 infected vaccinees of African-American descent. These results suggest that the immune stimulation provided by prior infections could partially overcome the limitations of genetic factors and strongly immunosuppressive treatments that are typically administered early after transplantation.

Nevertheless, these results clearly indicate that the transplant-related immunosuppressive regimens to which these patients were subjected did not inhibit the ability of the combined infection-vaccination stimulus to induce an effective neutralization response that extends to a relatively resistant virus, such as omicron. This is an encouraging result and suggests that adjuvant-based vaccines or other approaches that induce strong T-cell responses might be particularly beneficial for subjects with clinically induced immunosuppression.

## Data Availability

All data produced in the present study are available upon reasonable request to the corresponding author

## ACKNOWLEDGEMENTS

We thank the study participants for their generosity and willingness to participate in longitudinal COVID-19 research studies.

